# Resistant maltodextrin intake reduces virulent metabolites in the gut environment: randomized control study in a Japanese cohort

**DOI:** 10.1101/2020.05.25.20112508

**Authors:** Yuichiro Nishimoto, Yoshinori Mizuguchi, Yuka Mori, Masaki Ito, Shoko Miyazato, Yuka Kishimoto, Takuji Yamada, Shinji Fukuda

## Abstract

**Objective:** In recent years, there have been many reports on the effects of prebiotics on intestinal health. In particular, consuming resistant maltodextrin (RMD) has been reported to be beneficial. However, there has been no comprehensive quantification of the effect of RMD on the intestinal environment. Therefore, this study aimed to quantify the effects of RMD on the intestine, especially the intestinal microbiome and metabolome profiles.

**Design:** A randomized, double-blind controlled trial was conducted in 29 Japanese subjects with relatively high hemoglobin A1c (HbA1c). Subjects consumed RMD or placebo twice per day for 24 weeks. Blood and fecal samples were collected before and after intake. The intestinal environment was assessed by a metabologenomics approach combined with 16S rRNA gene-based microbiome and mass spectrometry-based metabologenomics analyses.

**Results:** The intake of RMD increased the levels of *Bifidobacterium* and *Fusicatenibacter*, and decreased deoxycholate. In addition, intake of the RMD lowered the levels of some virulent metabolites, such as imidazole propionate and trimethylamine, in subjects with an initially high amount of those metabolites.

**Conclusion:** RMD may have beneficial effects on the gut environment such as commensal microbiota modulation and reduction of virulence metabolites, known as a causative factor in metabolic disorders. However, its effect partially depends on the gut environmental baseline.

## Main Text

### Introduction

In recent years, there have been multiple reports that the intestinal microbiota plays a role in several diseases such as colorectal cancer, inflammatory bowel disease, obesity, and diabetes [1–4]. Additionally, the metabolites produced by the intestinal microbiota also affect human physiological homeostasis. For example, there have been reports on the effects of short-chain fatty acids (SCFAs) [5–7], vitamins [8,9], secondary bile acids [10], imidazole propionate (ImP) [11], and trimethylamine (TMA) [12–15] on human health and diseases. Thus, the gut environment has an impact on our health. One way to positively impact the intestinal environment is through the consumption of prebiotics. Resistant maltodextrin, also known as indigestible dextrin, is one of the dietary fibers that has been suggested to have prebiotic functions. The consumption of resistant maltodextrin (RMD) was found to increase the total bacterial number, particularly the genus *bifidobacteria* [16–18]. Additionally, it has been reported that SCFA levels were increased by the intake of RMD [16] but there are no comprehensive analyses of other metabolites. Moreover, the effect of diet, prebiotics, and probiotics on human health is different for each individual and based on their gut environment [19-21]. The purpose of this study was to observe the effects of the consumption of RMD on the intestinal environment through the following methods: i) metabologenomics analysis [22] of intestinal microbiota and metabolites before and after consumption of resistant maltodextrin; ii) a comparative analysis of microbiome and metabolome in individuals who presented with differences after consumption of resistant maltodextrin.

## Results

### Basic information on subjects and blood tests

To evaluate the effect of RMD, a randomized, placebo-controlled, parallel-group trial for 24 weeks was performed (Fig. 1a). A total of 30 Japanese participants with high hemoglobin A1c (HbA1c) levels passed the inclusion criteria, and 29 participants completed the intervention periods.

**Figure 1.**
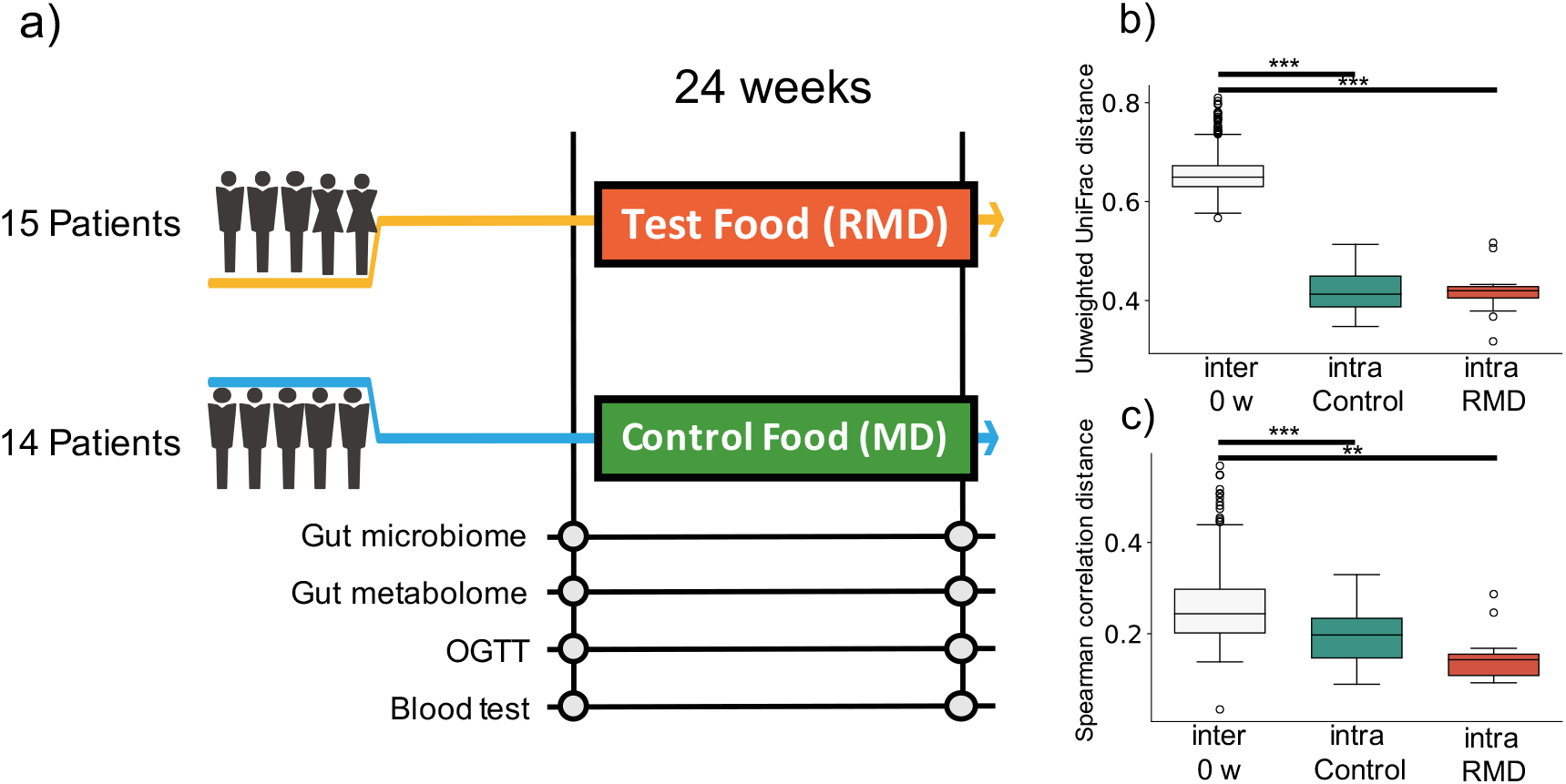
Experimental design of this study and comprehensive analysis of gut microbiome and metabolome profiles. (a): Flow diagram of the double-blind, placebo-controlled parallel group study. Fecal and blood samples were collected 0 and 24 weeks after test food (resistant maltodextrin: RMD) or control food (normal maltodextrin: MD) intervention. Gut microbiome and metabolome analyses, oral glucose tolerance test (OGTT), and blood test were conducted, (b): Box plot representing distribution of unweighted UniFrac distance for gut microbiome profiles among samples from different subjects at the same time point (inter 0 w) and the distance between samples from the same subject in control group and RMD group (*, *p <* 0.05; **, *p <* 0.005, ***, p < 0.0005; Dunn’s test), (c): Box plot representing distribution of the Spearman’s correlation distance for intestinal metabolome profiles among samples from different subjects at the same time point (inter 0 w) and the distance between samples from the same subject in control group and RMD group (*, *p <* 0.05; **, *p <* 0.005, ***, p < 0.0005; the Dunn’s test).

To investigate the benefits of RMD, parameters obtained in a clinical blood test, performed at two different time points, were evaluated with two hypothesis tests. There were significant decreases in insulin incremental area under the curve (iAUC) and area under the curve (AUC) as well as increases in blood glucose iAUC when compared with the baseline (q < 0.10). Items with significant differences before false discovery rate (FDR) correction have been described in Dataset S1.

### Effect of RMD on the intestinal microbiome and metabolome profiles

To evaluate the effect of RMD on the gut microbiota and the gut metabolic profile, we conducted fecal microbiome and metabolome profiling by metabologenomics approach. The result of a beta-diversity analysis showed that the microbiome and metabolome composition of the same subject were similar within the duration of the test food intake or placebo food intake. Comparisons of inter-individual, RMD-intra-individual, and maltodextrin (MD)-intra-individual distances showed significant differences (Fig. 1b and 1c; the Kruskal-Wallis test, p-values were 2.59×10^-18^ and 3.99×10^-8^ in the microbiota and metabolome data, respectively), suggesting that the individual difference was larger than the influence of the RMD or placebo food consumption.

Next, we performed two types of tests to evaluate the relative abundance of each microbiome and relative area of each metabolite. In the microbiome, the comparison between intra-group difference with the placebo group (see “Bioinformatics and Statistical Analysis” section in Methods) showed significant differences for some bacteria. However, there was no significant difference in the relative abundance of bacteria after FDR correction. We focused on the detected bacteria with significant differences when compared with the baseline. There were significant differences in five genera (*Bacteroides, Parabacteroides, Fusicatenibacter, Bifidobacterium*, and *[Ruminococcus] gauvreauii*) after FDR correction (Fig. 2, Dataset S1). In the metabolome analysis, there was no significant difference in metabolites after FDR correction in both tests. The significant differences before FDR correction in the two types of tests have been listed in Dataset S1. Noteworthy, the levels of deoxycholic acid and glycocholic acid were significantly decreased compared with those in the baseline (Fig. 3).

**Figure 2.**
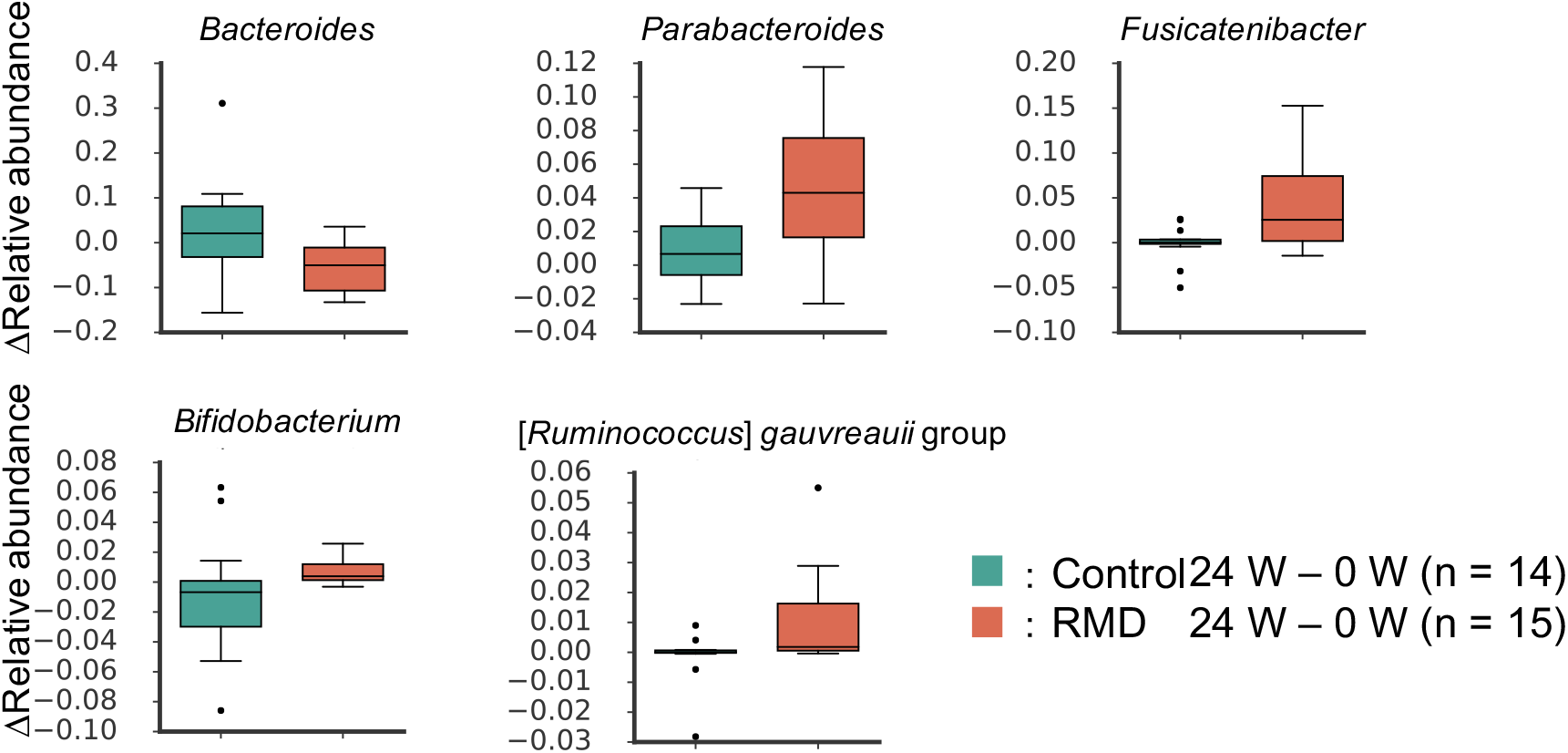
Significantly altered gut microbes by RMD intake. Box plots showing significant differences when compared to the control group and baseline. Box plots showing significant differences when compared to the control group (*p* < 0.05; the Wilcoxon rank sum test) and baseline (*q* < 0.10; the Wilcoxon signed-rank test).

**Figure 3.**
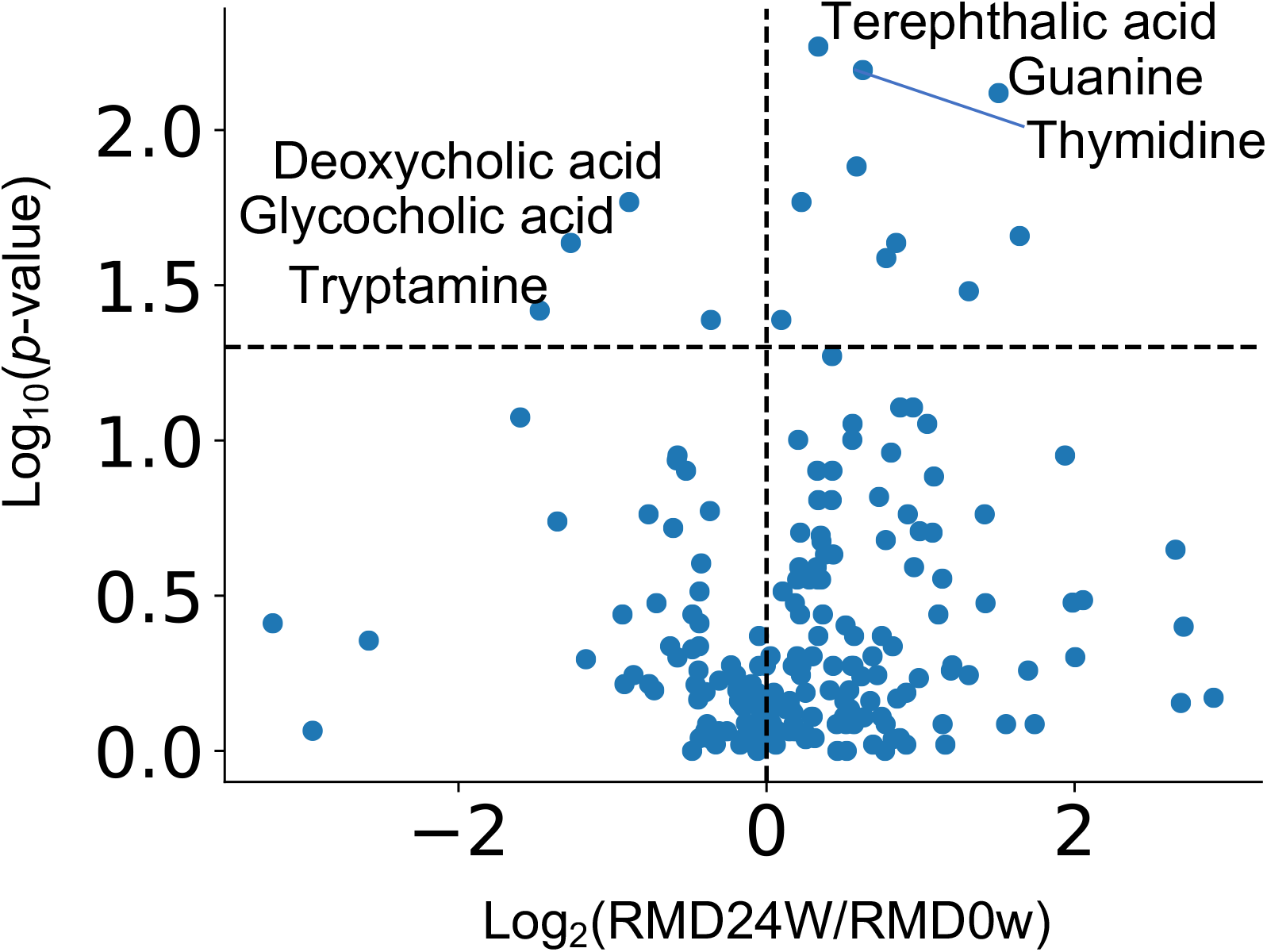
Effect of RMD intake on intestinal metabolite concentrations. Volcano plot representing the influence of the test food on each intestinal metabolite. X axis indicates the magnitude of influence (log fold change of mean value of corresponding metabolite abundance after 24 weeks of intervention relative to control time point). Y axis indicates the significance of influence (logarithmic value of p value). A dotted line has been drawn at -log10 p-value (*p* = 0.05).

### RMD intake alters each intestinal metabolite concentration depended on their baseline concentrations

It has been reported that the effects of diet, prebiotics, and probiotics on human health are individually different depending on the intestinal environment [19-21]. Therefore, we comprehensively analyzed the intestinal metabolites whose resulting variability depended on their baseline concentrations; 83 metabolites were selected including virulent metabolites (Fig. 4a; Dataset S2; q <0.10). This suggests that the effect of RMD intake on each intestinal metabolite concentration is different depending on the metabolite’s concentration at the baseline.

**Figure 4.**
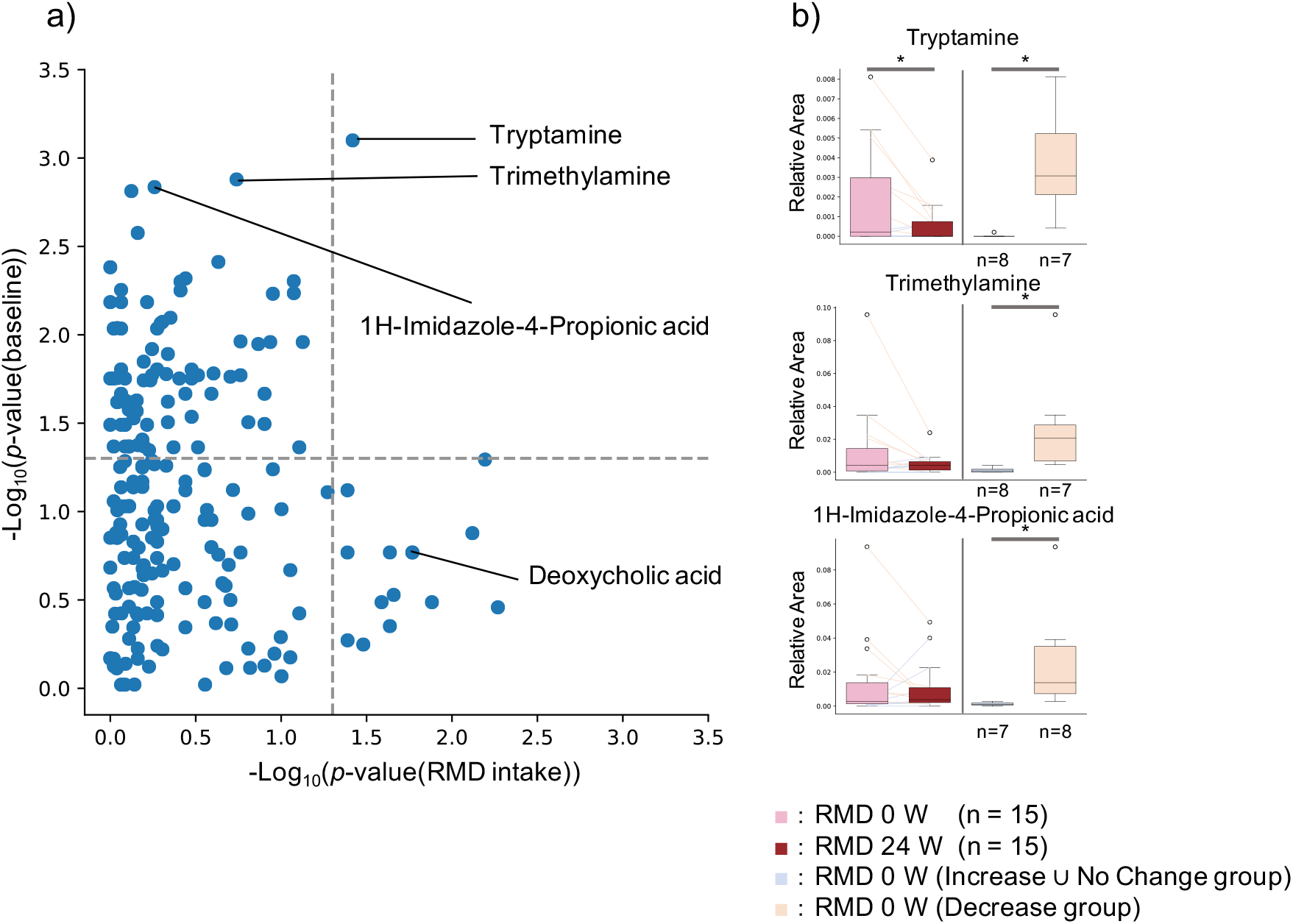
Effect of RMD intake on intestinal metabolite concentrations depended on the individual intestinal metabolite concentration itself at baseline. (a): X axis indicates the *P*-value of the Wilcoxon signed-rank test in each metabolite between RMD 0 w and RMD 24 w groups. Y axis indicates the *P*-value of the Wilcoxon rank sum test in each metabolite between 2 groups, increased and decreased groups after RMD intake, at the baseline. (If subjects with increased amounts of each metabolite were more than subjects with decreased amounts of each metabolite after RMD intake, Increase group and Decrease ⋃ No Change group were compared by the Mann-Whitney *U*-test; If subjects with decreased amounts of each metabolite were more than subjects with increased amounts of each metabolite, Decrease group and Increase ⋃ No Change group were compared by the Wilcoxon rank sum test.) (b): Three representative metabolites are shown by boxplots. The left two boxes show the amount of the metabolite in RMD 0 w (left) and RMD 24 w (right) (*, *p* < 0.05; the Wilcoxon signed-rank test), and the right two boxes show the amount of the metabolites at baseline in increase or comparative level of the target metabolites group (left) and reduction group (right) after RMD intake (*, *p* < 0.05; the Wilcoxon rank sum test).

### Intestinal environmental features in subjects who reduced virulent metabolite concentrations by RMD intake

ImP is metabolized from histidine by the gut microbiota and has been found to contribute to the pathophysiology of type 2 diabetes [11]. Our metabolome analysis showed that ImP levels after the RMD intake depended on the initial amount of ImP before RMD ingestion (Fig. 4b). To investigate the reason for the reduction of intestinal ImP, we analyzed the gut microbiome and metabolome by metabologenomics approach based on the following two criteria: (1) there was a significant difference of the gut microbial relative abundance and the relative amount of the metabolites during comparison of the subject who reduced ImP by RMD intake with the rest of the other subjects; (2) there was a significant difference between before and after RMD intake only for the subjects whose ImP decreased by RMD. There were no significant differences in the relative abundance of the gut microbiota in both analyses. However, some differences in metabolites such as ImP itself and TMA were detected as biomarkers for the reduction of ImP by RMD intake (Dataset S3). TMA is produced from dietary choline or carnitine by the gut microbiota and is metabolized to trimethylamine N-oxide (TMAO) in the liver. TMAO has been reported to increase the risk of arteriosclerosis [12,14]. Following the results of the same analysis for TMA, we found the intestinal microbes, such as *Desulfovibrio*, and metabolites, such as choline and TMA, could be used as biomarkers for TMA reduction by RMD intake (Dataset S4).

### Correlation analysis related to ImP and TMA

We found the two virulent metabolites, ImP and TMA were reduced in the subjects with a high amount of the two metabolites at the baseline. With an aim to explore the gut microbes responsible for the reduction in the levels of these two metabolites, we performed a correlation analyses between each microbial genus and ImP, or TMA (Dataset S5, S6). In the ImP, 6 genera (*Escherichia-Shigella, Allisonella, [Ruminococcus] gnavus* group, *Tyzzerella* 4, uncultured Lachnospiraceae, *Collinsella;* Spearman’s r > 0.38, *p* < 0.05) were found to have positive correlations and 10 genera (Ruminococcaceae UCG-013, *Phascolarctobacterium, Anaerostipes, Faecalibacterium, Ruminococcus 1*, Ruminococcaceae UCG-005, Lachnospiraceae NK4A136 group, uncultured Ruminococcaceae, Christensenellaceae R-7 group and Ruminococcaceae UCG-002; Spearman r < -0.36, *p* < 0.05) had negative correlations. In the TMA, 8 genera were found to have positive correlations (*Allisonella, Desulfovibrio*, Prevotellaceae NK3B31 group, *Ruminococcus 2, Clostridium* sensu stricto 1, *Megasphaera, Escherichia-Shigella* and uncultured Lachnospiraceae; Spearman’s r > 0.36; *p* < 0.05) and 3 genera (Ruminococcaceae UCG-002, Christensenellaceae R-7 group, and Ruminococcaceae UCG-005; Spearman’s r < -0.36, *p* < 0.05) had negative correlations. Interestingly, 6 genera such as *Escherichia-Shigella, Allisonella*, uncultured Lachnospiraceae, Christensenellaceae R-7 group, Ruminococcaceae UCG-002, and Ruminococcaceae UCG-005 were shared by both ImP and TMA, suggesting that these genera might have contributed to the reduction of the intestinal amount of ImP and TMA by RMD intake. Also, we noted that a positive correlation was found between ImP and TMA (Spearman’s r = 0.634, *p* = 0.000170).

## Discussion

There are various indicators that can be obtained from the measurement of blood glucose levels – from the indicators of blood glucose levels estimated at any point in time, to indicators of the blood glucose levels on the same day to those at a few or several months later (*e.g*. HbA1c, GA, 1,5-AG). Unexpectedly, HbA1c increased as a result of consumption of both RMD food and the placebo. It has been reported that HbA1c has seasonal patterns [23] thus the observed increment might have been affected by seasonal variations in this study. Regarding the glucose tolerance improvement, insulin AUC and iAUC decreased after the intake of RMD. However, the blood glucose iAUC increased significantly. This result is conflicting as it cannot explain the traditional relationship between decreasing insulin levels and improvement in the glucose tolerance. However, glycoalbumin (GA) and 1, 5-anhydroglucitol (1,5-AG) did not show significant differences. If glucose tolerance is impaired, an indicator that represents the blood glucose conditions, such as GA and 1,5-AG, will also deteriorate. These results show the limitation of the present study to conclude the effect of RMD on improvement in the glucose tolerance.

Some bacterial genera were significantly altered by the ingestion of RMD. Previous studies have reported that *Bifidobacterium* increase following an ingestion of RMD [18]. *Bifidobacterium* have been reported to produce mainly one SCFA, acetic acid, in the intestinal lumen. We previously reported that acetic acid produced by *bifidobacteria* upregulates the barrier function of intestinal epithelia to prevent Enterohemorrhagic *Escherichia coli* O157:H7 infection in a mouse model [24]. Thus, it is possible that increased numbers of *Bifidobacterium* may also upregulate the intestinal mucosal defense through acetic acid production in the human intestine. *Fusicatenibacter* has been known to degrade the polysaccharide inulin [25]. Thus, we hypothesized that *Fusicatenibacter* could metabolize RMD for nutrients for their growth in human intestine. Also, it has been shown that *Fusicatenibacter saccharivorans* is decreased in active ulcerative colitis patients, increased in quiescent ulcerative colitis patients, and suppresses intestinal inflammation through IL-10 secretion [26]. Thus, the increase of *Fusicatenibacter* after RMD intake may be valuable for suppression of the intestinal inflammation through IL-10 induction in the human intestine.

Although not significant after FDR correction, glycocholate and deoxycholate levels were reduced after RMD intake when compared to the baseline levels. Deoxycholate has been reported to trigger liver cancer when it is reabsorbed from the intestinal tract and reaches the liver [10]. Furthermore, we previously reported that the fecal concentration of deoxycholate was overrepresented in the early stages of colorectal cancer patient [1]. Therefore, the intake of RMD may reduce the risk of liver and colorectal cancer development. Bile acids promote micelle formation in the gastrointestinal (GI) tract. RMD has been reported to stabilize micelles [27] and thus may reduce the required amount of bile acid to produce deoxycholate by the gut microbiota in the GI tract.

Subjects who had decreased ImP levels in their feces after an RMD intake had significantly higher initial levels of ImP and TMA. Since ImP and TMA are positively correlated and some bacterial biomarkers for the reduction of them by RMD intake were shared, they may be controlled by similar mechanisms. In the ImP, the production may be stimulated by positively correlated bacteria (*Escherichia-Shigella, Allisonella, [Ruminococcus] gnavus* group, *Tyzzerella* 4, uncultured *Lachnospiraceae*, and *Collinsella*) and/or inhibited by negatively correlated bacteria (*Ruminococcaceae* UCG-013, *Phascolarctobacterium, Anaerostipes, Faecalibacterium, Ruminococcus 1, Ruminococcaceae* UCG-005, *Lachnospiraceae* NK4A136 group, uncultured *Ruminococcaceae, Christensenellaceae* R-7 group, and *Ruminococcaceae* UCG-002). It has been reported that ImP is produced by the gut microbiota from histidine via urocanate, and active sites of urocanate reductase (UrdA) must contain tyrosine or methionine [11]. In a report by Koh *et al*., it was shown that *Escherichia coli* K-12 had UrdA but the tyrosine and methionine were absent in its active site. The other four bacteria, *Allisonella, [Ruminococcus] gnavus* group, *Tyzzerella* 4, and *Collinsella*, have also been known to not express UrdA [11]. However, it has been reported that different strains of the gut microbiota have different functions, even in the same bacterial species [28]. Of these bacteria, an unreported strain may produce ImP. In addition, ImP decreased in subjects with high levels of ImP at the baseline. As ImP has been reported to impair insulin signaling through mechanistic target of rapamycin complex 1[11], subjects who have high levels of ImP in their feces may exhibit improved glucose tolerance through RMD intake.

Additionally, subjects who had reduced TMA levels after RMD intake had a significantly higher abundance of *Desulfovibrio*, and amount of choline and TMA at baseline. There are three possible pathways for TMA biosynthesis in the intestine: 1) the pathway undertaken by the dietary TMA in food to reach the intestines [29]; 2) the metabolism of dietary phosphatidylcholine/choline into TMA by the gut microbiota; and 3) the metabolism of dietary carnitine into TMA by the gut microbiota. It has been reported that phosphatidylcholine forms micelles with cholesterol. As shown in *in vitro* studies, RMD stabilized the micelles [27], thus RMD may reduce TMA production by inhibiting phosphatidylcholine hydrolysis. Subjects who have high levels of TMA in their feces may exhibit decreased arteriosclerosis risk through the intake of RMD.

A limitation of this study is that it is a parallel-group study. This report comprehensively analyzed and compared gut microbiome and metabolome profiles by metabologenomics approach among subjects who showed some effects after consuming RMD with those who did not. To more accurately evaluate the effects of RMD for each individual, a crossover study in which both RMD and placebo food are fed to each study participant will be necessary.

In this paper, a randomized control study was performed to quantify the effects of RMD on the intestinal environments using metabologenomics. As a result, we found that *Bifidobacterium*, a bacteria that promotes intestinal barrier functions through acetate production, was increased. *Fusicatenibacter*, which suppresses inflammation through IL-10, was also increased. Deoxycholate, which may cause liver and colorectal cancers, was decreased by the intake of RMD. In addition, it was found that if the amount of virulent metabolites RMD. Based on these findings, the pre-testing for such intestinal metabolite concentrations may allow for a personalized approach to improve the gut environment by RMD intake.

## Materials and Methods

### Trial Design and Recruitment

In this study, we conducted a randomized, double-blind, placebo-controlled, parallel-group clinical trial between September 2016 and February 2017. Another test period was set up as the target sample size was not reached in the first recruitment period (Fig. 1, Fig. S1, Dataset S7, S8, and S9). The baseline clinical characteristics were similar in both groups (Dataset S10). Each subject consumed test foods or placebo for 24 weeks. We used RMD as the test food and normal MD for the placebo. The RMD used in this trial was produced by Matsutani Chemical Industry Co., Ltd. (Itami, Japan; trade name: Fibersol-2; Dietary fiber: 90%). The calories in the test food and placebo were standardized in the trial although their weights were different; the test food and placebo were 10 g and 3 g, respectively. The test foods or placebo were dissolved in potable water and were consumed twice a day. The test food and placebo were individually packaged to prevent their recognition. Fecal sampling and oral glucose tolerance test (OGTT) were performed for each subject before the start of the study and at the end of the 24 weeks. Clinical blood tests were performed at the same time with OGTT. They included the measurement of HbA1c, blood glucose level, insulin, GA, 1,5-AG, glucagon-like peptide-1 (GLP-1), and tumor necrosis factor-alpha (TNF-α). For OGTT, the glucose and insulin concentrations were measured before administration and at 30 min, 60 min, 90 min, and 120 min after the administration of 75 g of glucose. The AUC and iAUC were calculated using the trapezoid model. This trial recruited males and females aged 20 years and older with high HbA1c levels (see Dataset S8 for detailed inclusion/exclusion criteria). Preliminary blood tests were conducted on the participants and 30 subjects who fulfilled the age, male-female ratio, and HbA1c level criteria from the preliminary results were selected. One subject (Subject 10) dropped out of this study as advised by a doctor; 29 subjects completed the main trial.

### Ethics approval

The human rights of the subjects who participated in this study were protected at all times, and the study observed the Helsinki Declaration and Ethical Guidelines on Epidemiological Research in Japan referring to cases concerning standards for clinical trials of drugs. This randomized controlled trial was conducted with approval of the clinical trial ethics review committee of the Chiyoda Paramedical Care Clinic (UMIN-CTR, Trial number: UMIN000023970).

### DNA Extraction

DNA extraction from stool samples was performed according to a previously described method [30]. From the extracted DNA samples, the V1-V2 region of the bacterial 16S rRNA gene region was amplified using universal primers 27F-mod and 338R [31]. DNA sequencing was performed with the DNA amplicons using the paired-end mode with 600 cycles (MiSeq, Illumina).

### Metabolite Extraction and Analysis

Metabolite extraction from feces was performed according to a previously described method [32]. Briefly, samples were lyophilized by using the VD-800R lyophilizer (TAITEC) for at least 24 h. Freeze-dried feces were disrupted with 3.0-mm zirconia beads by vigorous shaking (1,500 rpm for 10 min) using the Shake Master Neo (Bio Medical Science). Internal standards (20 μM each of methionine sulfone and D-camphor-10-sulfonic acid (CSA)) dissolved in 500 μl of methanol were added to 10 mg of the disrupted feces. Samples were further disrupted with 0.1-mm zirconia/silica beads by vigorous shaking (1,500 rpm for 5 min) using the Shake Master Neo. Next, 200 μl of ultrapure water and 500 μl of chloroform were added before centrifugation at 4,600 × *g* for 15 min at 20°C. Subsequently, 150 μl of the supernatant was transferred to a centrifugal filter tube (UltrafreeMC-PLHCC for Metabolome Analysis, Human Metabolome Technologies) to remove proteins and lipids. The filtrate was centrifugally concentrated and the pellet was resuspended in 50 μl of ultrapure water immediately before the capillary electrophoresis time-of-flight mass spectrometry (CE-TOFMS)-based metabolome analysis. The obtained metabolomic data has been shown in the Dataset S11 and S12.

### Bioinformatics and Statistical Analysis

The forward and reverse sequencing reads of each sample were merged using the vsearch version 1.9.3 (options: --fastq_maxee 9.0 --fastq_truncqual 7 --fastq_maxdiffs 300 -- fastq_maxmergelen 450 --fastq_minmergelen 250) [33]. Primer base nucleotides were removed by cutadapt (options: -O 13 -m 50 -M 450 -q 0; -e option is not used in cut 5’ primer, use 0.3 in cut 3’ primer). The Phix fragments were then removed using the Bowtie2 version 2.1.0 [34]. Bowtie2 option was set to default. Fragments with an average quality of less than 25 were removed by the in-house script. All fragments were mapped to the SILVA SSU Ref NR database version 128 [35] using Bowtie2 (Option: --no-hd --no-sq --no-unal -I 280 -X 400 --fr --no-discordant --phred33 -D 15 -R 10 -N 0 -L 22 -i S,1,1.15 -q). From the remaining fragments (26,784 ± 3,474), 10,000 fragments were subsampled and used for analysis.

All statistical analyses were performed using Python (version 3.7.3). The UniFrac distance were used for estimation of the beta diversity. The Wilcoxon-Mann-Whitney test was used to compare the two groups. False discovery rate was adjusted using the Benjamini-Hochberg false discovery rate correction (FDR-BH) method. For gut microbiome analysis, taxonomic composition data of the genus and the operational taxonomic unit (OTU) level were used. Of these, OTU level data was used only for calculation of beta diversities. The genus-level data was used for the other statistical analyses. For gut metabolome analysis, the relative area of metabolites data was used. The following comparisons were performed between the two different sets:

i. differences in the 24-week intake and baseline of each subject in the RMD and MD groups
ii. 24-weeks of RMD intake and baseline

Considering the placebo effect, test (i) was required. However, the effect of RMD on the intestinal bacteria and metabolites were smaller than that of the individual differences. Therefore, the effect of food may not be detected by test (i). Test (ii) was thus performed to increase the statistical significance of the test. As for microbiome data, we use only genera with an average relative abundance of 0.001 or more was used.

### Definition of Baseline-Dependent Metabolites

About each metabolite, subjects were divided into Increase (fold change > 1), Decrease (fold change < 1), and No Change groups (fold change = 1) by comparing results obtained before and after the RMD intake. For the initial value, (1) Increase group and Decrease ⋃ No Change group (2) Decrease group and Increase ⋃ No Change group were compared by the Mann-Whitney *U*-test. Items with a q value after FDR-BH correction of less than 0.10 were extracted. Since the α-error of the Mann-Whitney *U*-test becomes high if the proportions of the groups are different, the test was not performed if the number of people in a group was less than 5. In addition, when the not detected-sample is large, the behavior of its metabolite is unknown. Therefore, metabolites of which more than half were not detected were also excluded from this analysis.

### Data availability statement

The obtained 16S rRNA gene sequence data are available in the DDBJ DRA (DRA accession number: DRA010060). The obtained Blood test data has been shown in the Dataset S13.

## Data Availability

The obtained 16S rRNA gene sequence data are available in the DDBJ DRA (DRA accession number: DRA010060).

## Acknowledgments

The super-computing resource was provided by the Human Genome Center, the Institute of Medical Science, and the University of Tokyo. We would like to thank Matsutani Chemical Industry Co., Ltd. and CPCC Co., Ltd. who conducted the clinical trial. Additionally, we would like to thank the Metabologenomics, Inc. staff who participated in the discussion of this research.

## Author Contributions

Yoshinori M., S.M., Y.K., T.Y., and S.F. planned the study. Yuka M. and M.I. contributed to analysis of the primary sequence data of fecal samples. Y.N. contributed to the statistical analysis of the data. Y.N. wrote the first draft paper, Y.N., Yoshinori M., S.M., Y.K., T.Y., and S.F. contributed to the completion of the manuscript. All authors read and approved the final manuscript.

---

**Fig. S1.**
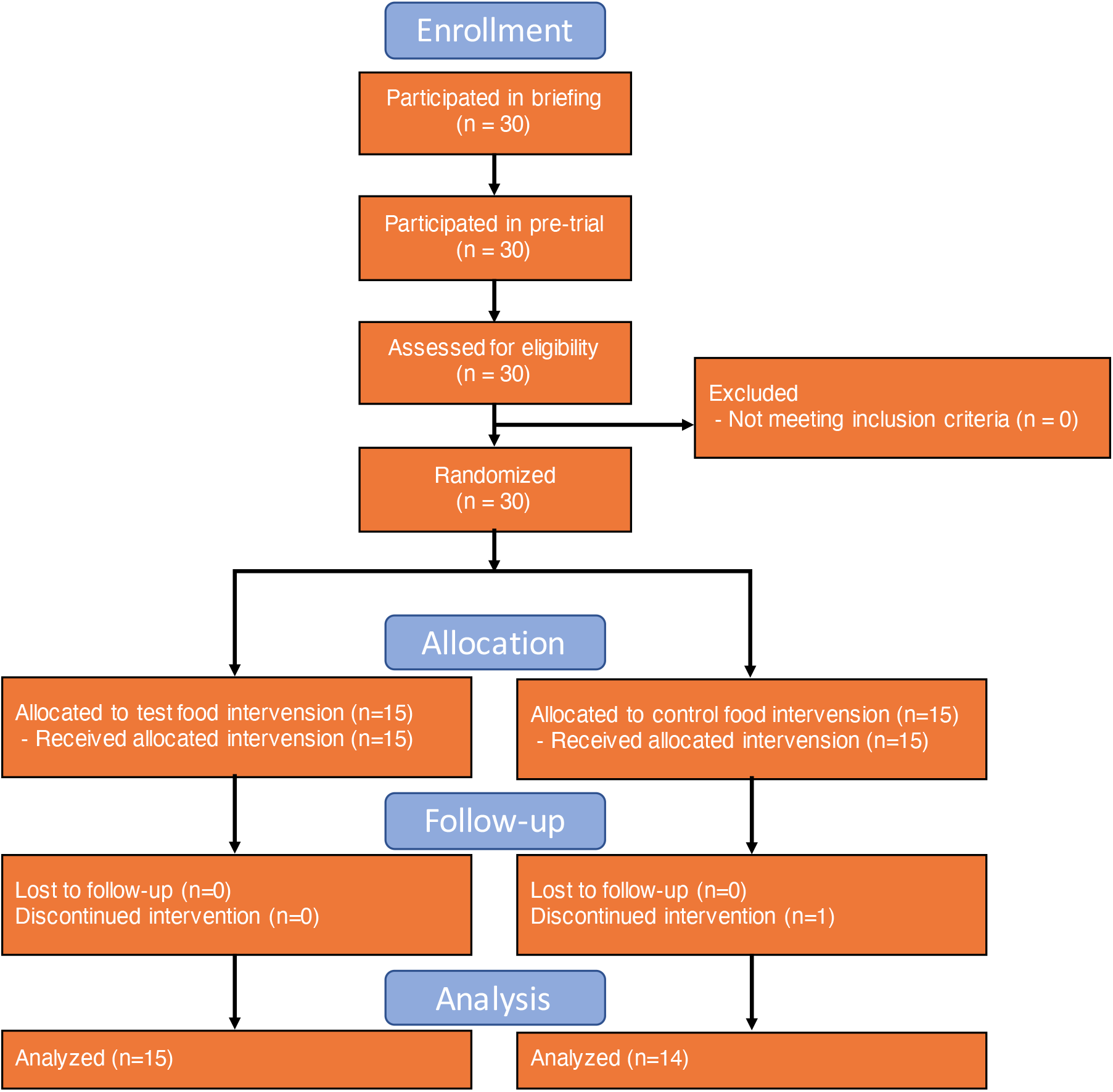
- Flow diagram of the phases of the randomised double-blind placebo-controlled parallel-group study.

